# Seroprevalence of anti-SARS-CoV-2 IgG Antibodies in the Staff of a Public School System in the Midwestern United States

**DOI:** 10.1101/2020.10.23.20218651

**Authors:** Lilah Lopez, Thao Nguyen, Graham Weber, Katlyn Kleimola, Megan Bereda, Yiling Liu, Emma K. Accorsi, Steven J. Skates, John P. Santa Maria, Kendal R. Smith, Mark Kalinich

**Affiliations:** Lake Central School Corporation, Saint John, IN; City of Evanston, Evanston, IL; Independent Researcher, Cleveland, OH; Biostatistics Center, Massachusetts General Hospital, Boston, MA; Harvard Medical School, Boston, MA; Department of Epidemiology, Harvard T.H. Chan School of Public Health, Boston, MA; Novartis Institutes for Biomedical Research, Cambridge, MA

**Author notes:** These authors contributed equally to the work (Lilah Lopez, Thao Nguyen, Graham Weber). Conflicts of interest: The authors declare they have no conflict of interest.

## Abstract

**Background:** Since March 2020, the United States has lost over 200,000 lives to severe acute respiratory syndrome coronavirus 2 (SARS-CoV-2), which causes COVID-19. A growing body of literature describes population-level SARS-CoV-2 exposure, but studies of antibody seroprevalence within school systems are critically lacking, hampering evidence-based discussions on school reopenings. The Lake Central School Corporation (LCSC), a public school system in suburban Indiana, USA, assessed SARS-CoV-2 seroprevalence in its staff and identified correlations between seropositivity and subjective histories and demographics.

**Methods:** This study is a cross-sectional, population-based analysis of the seroprevalence of SARS-CoV-2 in LCSC staff measured in July 2020. We tested for seroprevalence with the Abbott Alinity™ SARS-CoV-2 IgG antibody test. The primary outcome was the total seroprevalence of SARS-CoV-2, and secondary outcomes included trends of antibody presence in relation to baseline attributes.

**Findings:** 753 participants representative of the staff at large were enrolled. 22 participants (2·9%, 95% CI: 1·8% - 4·4%) tested positive for SARS-CoV-2 antibodies. Correcting for test performance parameters, the seroprevalence is estimated at 1·7% (90% Credible Interval: 0·27% - 3·3%). Multivariable logistic regression including mask wearing, travel history, symptom history, and contact history revealed a 48-fold increase in the odds of seropositivity if an individual previously tested positive for COVID-19 (OR: 48.2, 95% CI: 4 - 600). Amongst individuals with no previous positive test, exposure to a person diagnosed with COVID-19 increased the odds of seropositivity by 7-fold (OR: 6.5, 95% CI: 2.06 - 18.9).

**Interpretation:** Assuming the presence of antibodies is associated with immunity against SARS-CoV-2 infection, these results demonstrate a broad lack of herd immunity amongst the school corporation’s staff irrespective of employment role or location. Protective measures like contact tracing face coverings, and social distancing are therefore vital to maintaining the safety of both students and staff as the school year progresses.

**Funding:** Lake Central School Corporation

**Research in context:** *Evidence before this study:* We searched PubMed, SSRN, Research Square, and Gale Power Search for peer-reviewed articles, preprints, and research reports on the seroprevalence of anti-severe acute respiratory syndrome coronavirus 2 (SARS-CoV-2) IgG antibodies, published in English, using the search terms “COVID-19 in schools,” “COVID-19 seroprevalence,” “COVID antibodies,” and similar terms up to August 30, 2020. We identified several articles pertaining to the spread of COVID-19 within schools and among children. Current evidence on the pediatric transmission of COVID-19 is mixed, but early data on secondary school transmission are sobering. Shared among this literature was an acknowledgement of the paucity of data regarding how the pandemic may progress in the students and staff of primary and secondary education systems. To our knowledge, there is no study that specifically interrogates the seroprevalence of COVID-19 among US public school staff.

*Added value of this study:* As of September 2020, the United States has had more COVID-19 cases than any other country. With many US schools opening for in-person classes for the 2020-2021 school year, a granular understanding of the transmission dynamics within public school systems is vital to effectively and appropriately defending against COVID-19. Most seroprevalence studies have been based on city or hospital-level populations; this study establishes a baseline seroprevalence of SARS-CoV-2 antibodies in a Midwest public school district prior to the initiation of the school year.

*Implications of all available evidence:* The results of this study reveal that the majority (98·3%) of LCSC staff have not been exposed to COVID-19 prior to the start of the school year. Staff are therefore vulnerable to a large outbreak after the school opens, underscoring the importance of maintaining rigorous sanitary practices within the schools. It is vital that all members of LCSC and similar school districts across the country continue social distancing and mask wearing throughout the school day to limit exposure to COVID-19. Contact tracing in combination with rapid testing for individuals exposed to an individual with COVID-19 should also be employed.

## Background

Over 970,000 people around the world have lost their lives from severe acute respiratory syndrome coronavirus 2 (SARS-CoV-2, or COVID-19) as of September 2020. The United States alone has reported 200,000 deaths.^1^ A critical bottleneck in containing the virus is understanding its transmission dynamics. Although much effort has been expended on population-level seroprevalence surveys, scant data exist to understand COVID-19’s potential effect on the US public school system’s students and staff. Existing data are sobering: ten days into its reopening, an Israeli high school experienced a major outbreak.^2^ Work on the pediatric transmission of COVID-19 are conflicting; in multiple studies conducted in Southwest Germany, Ireland, and Northern France, children under 10 were found to have had little effect on the spread of the virus, while a study in Chile claimed that elementary students were more likely to contract the virus relative to secondary students.^3,4,5,6^ Here, we determine the seroprevalence of COVID-19 in the Lake Central School Corporation (LCSC), a public school system located in suburban Indiana, US. Although there is enormous heterogeneity within the US public school system, LCSC staff’s demographics are broadly representative of national statistics. The LCSC has 16·6 students per teacher, similar to the United States average of 16 students per teacher.^7,8^ The median age of public school employees in the United States is 41, while the median age of the Lake Central employee population lies at 48.^9,10^ Notably, previous reports have established that advanced age is highly associated with COVID-19 hospitalization, further underscoring the threat of morbidity and mortality within this community relative to other school districts.^11^ Given the lack of specificity of COVID-19 symptoms and that mild and asymptomatic cases of COVID-19 may go undocumented, antibody-based seroprevalance studies are required to estimate population-level exposure to SARS-CoV-2, although it should be noted that immunity to SARS-CoV-2 via such antibodies has yet to be firmly established.^12^

## Methods

### Study design and participants

This cross-sectional, population-based analysis of the seroprevalence of anti-SARS-CoV-2 IgG enrolled participants over a five day period in July of 2020 during participation in the LCSC’s annual wellness check. Individuals were eligible to participate if they were 18 years of age or older and were employees of the Lake Central school corporation during the 2018-2019 or 2019-2020 school years.

After approval from the Community Healthcare System Central Institutional Review Board (CHS CIRB #07-02), participants were contacted through their respective LCSC email account as well as by voice message from the LCSC superintendent informing them of the opportunity to participate. These communications provided information regarding the study and a link in the email allowed participants to schedule their testing date. A second email containing a video promoting the study was sent to staff and shared on LCSC social media accounts. There was no cost to participate in testing. Once registered, each participant completed a data questionnaire containing questions about sociodemographic characteristics including self-identified gender, employment factors, and activities that can increase the risk of having COVID-19, as well as informed written consent (Supplementary Materials).

During the five days of data collection, the study team was present at the testing site to assist with distributing additional consent forms and data questionnaires. Participants were required to wear a mask upon entering the building and hand sanitizer was available at each station.

Stations were six feet apart, and a member of the LCSC staff used cleaning wipes to disinfect each station between participants. An independent third party vendor, Franciscan WorkingWell, performed venipuncture using standard procedure for the antibody tests, and obtained a 5 mL blood sample for the antibody test and a 6 mL blood sample for blood typing.

The primary outcome was the total seroprevalence of anti-SARS-CoV-2 IgG antibodies within staff. The secondary outcomes were the changes in odds of seropositivity associated with baseline demographics and COVID-19 related factors including mask use, contact history, self-reported exposure to a known COVID-19 positive person or persons, and a previous positive COVID-19 test (PCR or antibody-based) (Supplementary Materials).

### Laboratory analysis

Seroprevalence was determined utilizing the commercially available Abbott Alinity™ SARS-CoV-2 IgG antibody test, with a reported 100% sensitivity (34/34, 95% CI: 89·7%-100%) and 99% specificity (99/100, 95% CI: 94·5%-100%) in detecting anti-SARS-CoV-2 IgG antibodies.^13^ This test provides a binary, present/not present result.

### Statistical analysis

Seroprevalence among LCSC staff was calculated as the proportion of staff members who received a positive antibody test out of the total staff tested. Confidence intervals were first estimated by generating a binomial confidence interval with the statsmodel package in Python v3.7. The seroprevalence was then corrected for uncertainties in the test sensitivity and specificity using previously established statistical approaches.^14^ Relative risks (risk ratios) were calculated to identify associations between seropositivity and gender, BMI, blood type, contact history, symptom history, travel history, role in school corporation, school where employed, extracurricular role (coach, club sponsor), mask history, previous positive COVID-19 test, and age on 7/20/20, the last day of sample collection. Employment role was separated into two groups: those departments that worked in the school during the Indiana stay-at-home order (maintenance, technology, and administration) and those that did not (all others). Continuous variables (BMI, age) were binarized by segmenting the variable by its median value. Bonferroni correction was used to correct for multiple testing on the univariate analyses. To determine if having a history of mask-wearing reduced seropositivity, we constructed a directed acyclic graph (DAG) based on existing literature evidence and identified gender, age, travel history, school type, and role at LCSC as potential confounders to control for in the multivariable logistic regression (Supplementary Figure 1).^6,15^ Finally, backward-stepwise feature elimination was used to identify the features most predictive of seropositivity in a multivariable logistic regression starting with the thirteen categories queried in the survey (threshold *p*-value of 0·05). After the model was identified for which all factors were statistically significant, three factors (mask history, travel history, and symptom history) which are understood based on external studies to have an impact on infectivity were reintroduced to this reduced model.^13,14,15^

One factor, having a previous positive COVID-19 test in four individuals, was overwhelmingly predictive for seropositivity. The regression was then re-fitted on the vast majority of individuals without a previous positive test to estimate associations of features with seropositivity within this group. For continuous variables such as age and BMI, missing data were either removed (univariate analyses) or replaced with the median of existing data for that variable (multivariable analysis). Missing data for categorical variables were either removed (univariate analyses) or replaced with the mode of that variable (multivariable analysis). Logistic regressions were calculated using the scikit-learn Python package.

## Results

Of the eligible 1261 staff members, 753 (60%) participated in the study. The LCSC staff comprises 1060 (84%) women; the participation group had 635 (85%) women; age demographics for the study population compared to that of the total staff were similarly representative (Table 1).

**Table 1.** Characteristics of Study Population and Total Staff Population.

| Characteristic | Study Population |  | Total Staff Population |  |
| --- | --- | --- | --- | --- |
|  | *n* | % | *n* | % |
| Gender* |  |  |  |  |
| Female | 635 | 84.7 | 1060 | 84.1 |
| Age** |  |  |  |  |
| 18-35 | 153 | 20.3 | 261 | 20.7 |
| 36-50 | 287 | 38.2 | 483 | 38.3 |
| 51-65 | 282 | 37.5 | 455 | 36.1 |
| 66+ | 30 | 4.0 | 62 | 4.9 |

*Note: Three participants did not share their gender and one did not share their age.*
*\*Chi-square value 0.0025, p-value 0.960.*
*\*\*Chi-square value 1.308, p-value 0.727.*

22 individuals tested positive for anti-SARS-CoV-2 IgG antibodies, representing 2·9% seroprevalence (95% CI: 1·8% - 4·4%). After correcting for the reported sensitivity and specificity of the IgG antibody test, the seroprevalence is estimated as 1·7% (90% Credible Interval: 0·27% - 3·3%).^16^

In the univariate analyses, having a previous positive COVID-19 test (RR 29·5, 95% CI: 14·3 - 60·4, p<0·0001) or contact with a COVID-19 case (RR 6·86, 95% CI: 3·04 - 15·5), p<0·0001) were found to be statistically significantly associated with seropositivity, after Bonferroni correction (Table 2). No causal relationship between self-reported mask-wearing history and seropositivity could be identified, after controlling for six confounders available in our data set, identified using a directed acyclic graph (Supplementary Table 1) (RR 0.83, 95% CI 0.18 - 3.8). In the multivariable model employing backward-stepwise feature elimination, a previous positive COVID-19 test (OR 48·2, 95% CI 3.9 - 597, p=0·003) or contact with a COVID-19 case (OR 5·6, 95% CI 2.1 - 15.1, p=0·001) were found to be the most predictive factors for seropositivity (Supplementary Table 2). To interrogate the specific interaction between contact history and seropositivity, the dataset was subsetted to the vast majority of individuals without a previous positive COVID-19 test (n=749). In this population, a logistic regression analysis including the predictors mask wearing, travel history, and symptom history, a positive contact history conferred a 7-fold increase in the odds of seropositivity (p<0·0001) (Table 3). The odds increased from 1:40 without such a contact to 1:5.5 with contact - or the probability of seropositivity increased from 2.5% to 15%.

**Table 2.** Univariate Analysis.

| Characteristic | Relative Risk | 95% Confidence Interval | $p$ |
| --- | --- | --- | --- |
| Covid Status (Self-reported Previous Positive Test) | 29.5 | (14.3, 60.4) | $< 0.0001$ |
| Contact History (Yes) | 6.86 | (3.04, 15.5) | $< 0.0001$ |
| Club Sponsor | 3.10 | (1.22, 7.84) | 0.0258 |
| Symptom History (Yes) | 2.31 | (0.990, 5.42) | 0.0577 |
| BMI (Binary) | 2.17 | (0.884, 5.25) | 0.0867 |
| Was Working at school (in the summer) | 0.196 | (0.027, 1.45) | 0.0979 |
| Sports Coach | 1.96 | (0.591, 6.49) | 0.225 |
| Gender (Female) | 0.616 | (0.232, 1.64) | 0.362 |
| Mask Wearing (Yes) | 0.721 | (0.172, 3.03) | 0.654 |
| Age (Binary) | 1.28 | (0.559, 2.92) | 0.667 |
| Travel History (Yes) | 0.744 | (0.255, 2.17) | 0.798 |
| Elementary School | *ref* | - |  |
| Middle School | 2.25 | (0.813, 6.20) | 0.117 |
| High School | 1.15 | (0.357, 3.73) | 1.00 |
| Other School | 1.10 | (0.226, 5.35) | 1.00 |
| Blood Type (O+) | *ref* | - |  |
| Blood Type (B-) | 3.28 | (0.475, 22.6) | 0.286 |
| Blood Type (B+) | 0.336 | (0.044, 2.58) | 0.468 |
| Blood Type (A+) | 0.758 | (0.293, 1.96) | 0.628 |
| Blood Type (A-) | 0.444 | (0.058, 3.40) | 0.696 |

AB+, AB-, and O- had no positive cases and therefore have no RR estimate.

**Table 3.** Multivariable Model.

| Effect | Odds Ratio | 95% CI |  | *p* |
| --- | --- | --- | --- | --- |
|  |  | *LL* | *UL* |  |
| Intercept | 0.029 | 0.0044 | 0.10 | <0.001 |
| Contact History | 6.5 | 2.06 | 18.9 | <0.001 |
| Mask History | 0.58 | 0.15 | 3.87 | 0.49 |
| Travel History | 0.36 | 0.05 | 1.33 | 0.18 |
| Symptom History | 1.62 | 0.52 | 4.57 | 0.38 |

Note: CI = confidence interval; *LL* = CI lower limit; *UL* = CI upper limit.

## Discussion

This study provides the necessary baseline for future longitudinal monitoring to assess the impact of public school reopening on COVID-19 transmission. With a demographically representative 60% staff participation rate, these data offer an unprecedented view into the seroprevalence of the target population of a Midwest US public school system. The estimated 1·7% seroprevalence of COVID-19 antibodies among LCSC staff is far below that reported in metropolitan areas throughout the US at the time of testing, underscoring the large and continued risk of COVID-19 transmission within this community upon exposure.^16^ As a point of comparison, Lake County, Indiana, the county within which the LCSC resides, had 4,985 reported cases of COVID-19, representing 1.0% of the county’s population.^17^

Although the effect of mask wearing on seropositivity, utilizing a directed acyclic graph to identify potential confounders, did not achieve statistical significance, this result should not be interpreted as a lack of efficacy in the use of masks given the overwhelming quality and quantity of data supporting their continued use, and is likely due to lack of power.^18^ Both univariate and multivariable analyses demonstrate that a previous positive COVID-19 test or a history of contact with a COVID-19 patient were associated with seropositivity. The 7-fold increase in odds of seropositivity for individuals with a positive contact history is especially noteworthy amongst the vast majority (n=749) of the individuals without a previous positive COVID-19 test. Although these relationships are associational, the combination of low staff seroprevalence and the strong positive association of seropositivity with contact history highlights a high risk group for which the importance of aggressive contact tracing would be efficient and which would minimize the transmission of COVID-19, as well as continued protective procedures during school hours such as mask and face shield wearing, social distancing, and frequent disinfection of school property.^19^

This study has several limitations. Although data were collected from staff employed at 11 different sites (10 schools and the transportation facility), only the LCSC was involved in the study, limiting the generalizability of this work. Neither ethnicity nor income data were collected, precluding analysis of these variables’ previously demonstrated associations with COVID-19 positivity.^20^ We rely on study participants to self-report variables, excluding antibody status and blood type, via questionnaire. Some questions may be insufficiently granular, such as the binary mask wearing variable, and participants may also make errors filling out the questionnaire.

Insufficient granularity is likely to make groups defined by the variable more similar to each other, while reporting errors are likely to be random and unrelated to the outcome, antibody status. Thereby both will be expected to create bias towards the null, meaning that the results reported in this study are conservative. The low baseline seroprevalence (22 positive tests in the 753-person cohort) prevents the identification of more subtle, but potentially real, associations among the collected variables and seropositivity. Additionally, the lack of county-level seroprevalence data prevents comparison to the broader population outside of the school district. Finally, data was not collected on students in the corporation, preventing the investigation of a potential link between staff or student positivity and transmission within or between these two co-exposed groups.

The reopening of US schools poses a potential risk of COVID-19 transmission to both staff and students.The low pre-opening seroprevalence, in combination with the advanced age of a significant fraction of the LCSC staff, may increase the number and severity of cases should an outbreak occur within this school system, or other school systems sharing its demographic characteristics. Teachers should consider deploying novel teaching strategies that limit the amount of non-distanced interactions. Administrators and legislators should allocate the required resources to implement and maintain robust personal protective measures, including contact-tracing, masks, face shields, and social distancing to protect the lives of both the children of the school district and those charged with educating them.

## Data Availability

Limited data are available to researchers upon reasonable request for data sharing to the corresponding author after IRB approval.

## Contributors

KRS, LL, TN, GW, and MK conceived the study. LL, TN, and GW drafted the first version of the manuscript. MK, MB, KK, EA, and KRS contributed to drafting sections of the manuscript. LL, TN, GW, MK, KK, YL, SJS, YL, EA, and JSM performed data analyses. All authors contributed to the interpretation of data and read and approved the final manuscript.

## Acknowledgements

We thank all of the LCSC staff members who participated in this study. We also thank Jana L. Lacera, Director, IRB/Bio-Ethics, who graciously volunteered both her and her organization’s resources; the nurses from Franciscan WorkingWell for collecting participant samples; Dr. Marc Lipsitch for his invaluable guidance, feedback and encouragement; and the LCSC for their generosity in funding this work.

## Supplementary Materials

Data Collection for Seroprevalence Study of COVID 19 in Lake Central Staff

**This is a student-led study with the purpose of determining the number of Lake Central staff members who have been exposed to COVID 19. If you do not wish to participate in this study, your action on this form is not necessary**.

Print: ________________

1. Date of Birth (MM/DD/YYYY): __ __ / __ __ / __ __ __ __
2. Height (ft): ________
3. Weight (lb): ________
4. Gender: Male □ Female □ Other □ Prefer not to answer □
5. a. Have you shown any of the following symptoms (check all that apply):
  - Headache □
  - Cough □
  - Congestion □
  - Nausea □
  - Loss of taste/smell □
  - Fever or Chills □
  - Fatigue □
  - Shortness of breath/difficulty breathing □
  - Muscle/body aches □
  - Diarrhea □
  - Sore throat □
b. If yes, when did you last experience those symptoms (MM/DD): __ __ / __ __
6. a. Have you been in contact with anyone who has been diagnosed with COVID-19: Yes □ No □
b. If yes, when (MM/DD): __ __ / __ __
7. a. Have you travelled outside the state in the past two weeks: Yes □ No □
b. If yes, to where (City, State): ___________________
8. a. Have you tested positive for COVID-19: Yes □ No □
b. If yes, when did you test positive (MM/DD): __ __ / __ __
9. Do you wear a mask while out in public: Yes □ No □
10. If you are not a Lake Central Staff Member

a. What is your relation to the staff member: __________________
b. What is the name of the staff member: __________________ **If you are not employed by Lake Central School Corporation, please refrain from answering questions 10-13**.
11. Role at LC (Check one):
  - Janitorial/Maintenance Staff □
  - Technology Department □
  - Lunch Staff □
  - Bus Driver/ Bus Aide □
  - Guidance/Administration □
  - West Lake Department □
  - World Language, Business, Consumer Science, or Communications Department □
  - General Elementary Teacher □
  - Art, Music, or P.E. Department □
  - English Department □
  - Math Department □
  - Science Department □
  - Social Studies Department □
12. What school do you work at (Check all that apply):
  - Lake Central High School □
  - Clark Middle School □
  - Kahler Middle School □
  - Grimmer Middle School □
  - Watson Elementary School □
  - Homan Elementary School □
  - Peifer Elementary School □
  - Kolling Elementary School □
  - Bibich Elementary School □
  - Protsman Elementary School □
  - Transportation Center □
13. Do you coach a school sport: Yes □ No □
14. Do you supervise a school club: Yes □ No □

**Supplementary Figure 1:**
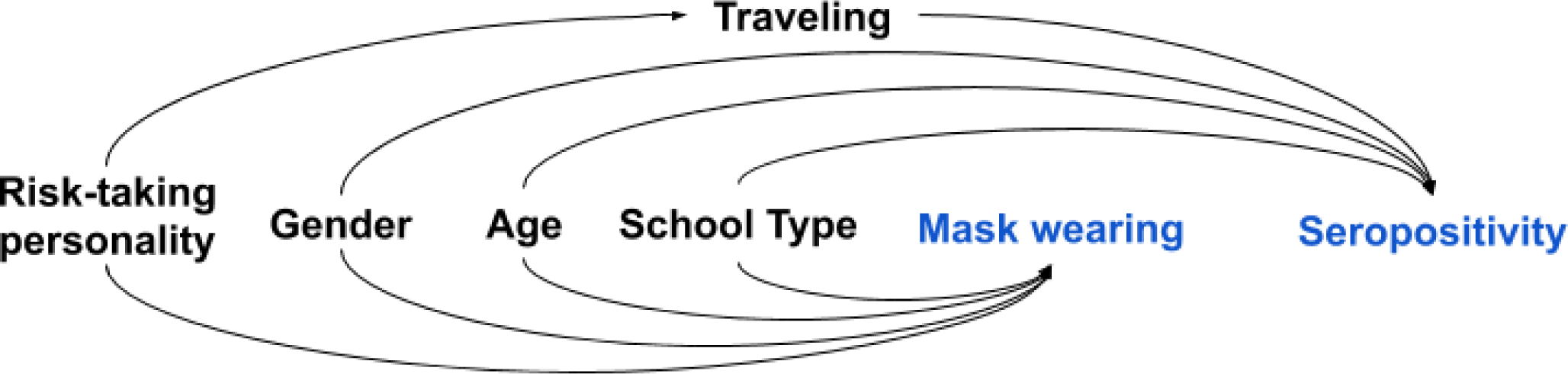
Construction of a DAG to Test for a Causal Association between Mask Wearing and Seropositivity. *Risk-taking personality could affect seroprevalence in other ways such as not social distancing, but those data weren’t collected nor adjusted for in this round of testing.

**Supplementary Table 1:** Logistic Regression Results for the relationship between mask wearing history and seropositivity, adjusting for potential confounders.

| Effect | Relative Risk<br>Ratio | 95% CI |  | *p* |
| --- | --- | --- | --- | --- |
|  |  | *LL* | *UL* |  |
| Constant | 0.028 | 0.0015 | 0.51 | 0.016 |
| Working in the Summer | 0.16 | 0.022 | 1.3 | 0.082 |
| Working in a Middle School | 3.4 | 0.69 | 17 | 0.13 |
| Female Gender | 0.56 | 0.19 | 1.6 | 0.29 |
| Travel History | 0.66 | 0.22 | 2.0 | 0.47 |
| Working in a High School | 1.7 | 0.31 | 9.1 | 0.55 |
| Age | 1.0 | 0.97 | 1.0 | 0.67 |
| Working in Elementary School | 1.4 | 0.26 | 7.5 | 0.69 |
| Mask History | 0.83 | 0.18 | 3.8 | 0.82 |

**Supplementary Table 2:** Stepwise Backwards Feature Elimination Regression Results.

| Effect | Odds Ratio | 95% CI |  | *p* |
| --- | --- | --- | --- | --- |
|  |  | *LL* | *UL* |  |
| Intercept | 0.028 | 0.0063 | 0.13 | <0.001 |
| Contact History | 5.6 | 2.1 | 15.1 | 0.001 |
| Previous COVID Test | 48.2 | 3.9 | 597 | 0.003 |
| Mask History | 0.61 | 0.13 | 2.8 | 0.53 |
| Travel History | 0.66 | 0.21 | 2.1 | 0.48 |
| Symptom History | 1.6 | 0.61 | 4.4 | 0.33 |

